# National interest may require distributing COVID-19 vaccines to other countries

**DOI:** 10.1101/2021.01.11.21249610

**Authors:** Tiziano Rotesi, Paolo Pin, Maria Cucciniello, Amyn A. Malik, Elliott E. Paintsil, Scott E. Bokemper, Kathryn Willebrand, Gregory A. Huber, Alessia Melegaro, Saad B. Omer

## Abstract

As the clinical trials for COVID-19 vaccine progress, understanding how to distribute the initially scarce doses is of paramount importance and a quantitative analysis of the trade-offs involved in domestic-only versus cooperative distribution is still missing. In this study we use a network Susceptible-Infected-Removed (SIR) model to show under which circumstances it is in a country’s self-interest to ensure other countries can also obtain a COVID-19 vaccine rather than focusing only on vaccination of their own residents. In particular, we focus our analysis on the US’s decision and estimate the internal burden of COVID-19 disease under different scenarios about vaccine cooperation. We show that in scenarios in which the US has reached the threshold for domestic herd immunity, the US may find it optimal to donate doses to other countries with lower vaccination coverage, as this would allow for a sharp reduction in the inflow of infected individuals from abroad.

## Main text

The COVID-19 pandemic has caused enormous morbidity and mortality globally with over 75 million cases and almost 1.7 million deaths as of December 2020^1^. In the absence of a vaccine or substantively effective pharmacological interventions, governments have instituted stay-at-home orders and border lockdowns to slow the spread of the disease^2^. However, this has resulted in disruption of many economic activities with resulting financial losses and long-term consequences^3-4^. An effective vaccine that allows societies to achieve sufficient immunity to stop the exponential spread of the disease is therefore widely seen as necessary for a speedy return to pre-pandemic practices. As of January 7, 2021, there are 64 vaccines in clinical trials on humans, with 3 already approved and 20 in phase 3 trials^5^.

As the first vaccine has been approved both in the UK and in the *US*, scientists have turned to the key question of how the initially scarce vaccine will be distributed. While a great deal of this focus has been on prioritization *within* countries, there is a perhaps even more important question about how to allocate the vaccine *across* countries. Many countries are taking steps to ensure that they would have adequate supplies of the vaccine for their population. But, the optimal strategy for securing vaccine supplies and balancing the global-interest with individual countries’ self-interest is unclear. As the discussion on equitable vaccine distribution continues, some countries have already reserved significant doses of vaccines for themselves^6^.

The World Health Organization launched the COVAX pillar of the Access to COVID-19 Tools Accelerator to provide a global solution and equitable access to vaccines^7^. COVAX Facility aims to support the development and manufacturing of various COVID-19 vaccine candidates and negotiate pricing. The COVAX Advance Market Commitment (AMC) is an important component of COVAX Facility that will provide vaccines to 92 low and lower-middle income countries through a unique funding mechanism leveraging the required volume and buy-in from higher income countries participating in COVAX Facility. However, not all countries are participating in COVAX Facility. A few countries have struck only individual deals with the manufacturers most likely to have an effective vaccine, resulting in concerns about ‘vaccine nationalism’^8^. This country-by-country approach ignores the fact that ending a global pandemic requires all countries to have sufficient access to the vaccine for their populations.

Because closing international borders^9^ over the long-term is likely not an economically viable option^10-12^, it may be in the self-interest of countries that have greater access to the vaccine to cooperate and share the vaccine with other. The home country-first approach also carries a risk that if a vaccine manufacturer is not successful, countries signing deals only with them may be locked out of a vaccine. However, whether, and under which conditions, it is in a resource rich country’s interest to support vaccine allocation to other countries is not currently known.

## The model

To study the effect of cooperation actions versus uncooperative vaccine distribution behavior, we use a network model where countries are nodes and links are individuals moving from one country to another. For each country, a separate compartmental SIR model was constructed, whereby individuals can become infected through interaction with SARS-Cov-2 positive cases under the assumption of internal homogeneous mixing. Network effects are such that the safety of one country also depends on the risk of having large shocks in other parts of the world, especially in those countries where herd immunity levels have not been achieved. Specifically, if there is an outbreak in one country, infected individuals may spread it to other countries through the modeled mobility patterns. We model international travel using data on air travel and cross-border ground travel. Airline transportation data gathered by the International Civil Aviation Organization (ICAO) allows us to estimate the average daily number of passengers from each country of origin to the rest of the network. Ground mobility flows from the US to both Mexico and Canada, and *vice versa*, are used to estimate those travel patterns (Fig S1 in the supplementary material represents the mobility network).

In a standard SIR framework, herd immunity is achieved when the share of protected individuals is above a certain threshold *P*_*c*_ = 1 − 1/*R*_0_^13^. The higher the average number of secondary cases that can be generated by one single infectious case (*R*_0_), the larger the proportion of individuals in a population who must be vaccinated to reach the herd immunity level. This is particularly relevant when studying the number of individuals who become infected in the presence of a single primary case (Fig 1). When one infectious person appears in a given population, the number of secondary cases changes as a function of how many in the population are susceptible. When a country is close to the threshold of herd immunity, there is a clear discontinuity in the number of cases that follow from a new infection, as the effect of a shock increases by several orders of magnitude once herd immunity is lost.

**Fig. 1.**
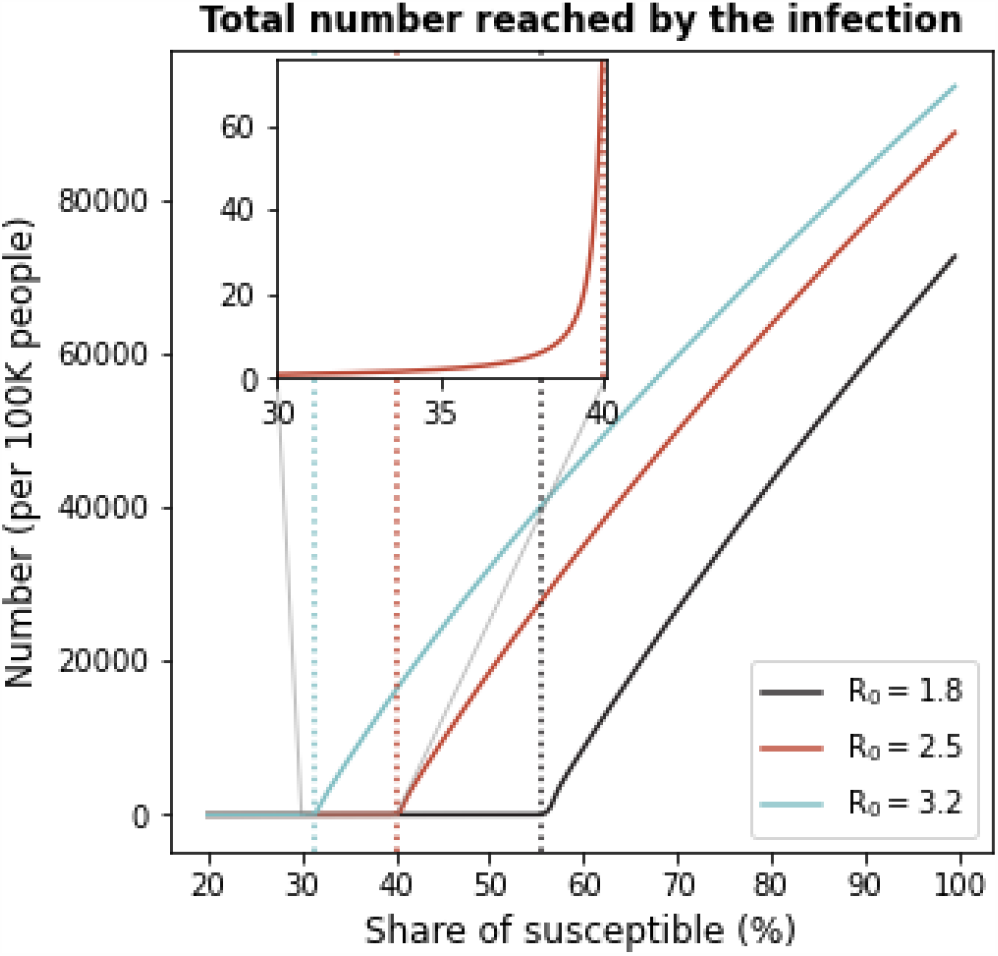
Number of infected individuals as a function of the share of susceptible in the population. When the proportion of susceptible individuals crosses the herd immunity threshold (1/*R*_0_), the number of infections increases drastically. Dotted lines represent herd immunity levels associated with three levels of *R*_0_.

We start by defining a measure of risk, which captures the degree of exposure of each country to infections from the rest of the network (see section 1 in the supplementary material) and we find that the US is particularly exposed (Fig S2). Focusing on the US, we performed simulations assuming that an infection starts somewhere else in the world. This scenario is representative of a situation in which a population with low immunity levels experiences an internal outbreak that may then travel to other countries through individuals’ mobility patterns. We then estimate how many people we expect to get infected in the US.

### Two vaccine allocation strategies

Two counterfactual alternative vaccines allocation strategies are evaluated. One is a domestic- only allocation strategy and in the other the US donates some doses of its vaccine stock to other countries. Specifically, we assume that the US has available a number D of extra doses of a vaccine with a modeled efficacy of 95%^14^. These doses could either be kept for domestic distribution (uncooperative strategy) or else distributed in any other country (cooperative action) eligible for COVAX AMC. To compare the output of the cooperative and the uncooperative action, we run simulations of our model and estimate the difference in the overall number of domestic cases in the US. The framework of our analysis can be applied to an initial period of vaccine allocation, but also to a later condition in which a country has allocated some doses domestically and is getting D additional doses.

We note that this is a hard test of the claim that cooperative vaccine distribution is desirable. While sharing a scarce vaccine stock with other countries will reduce deaths in those receiving countries, we ignore those welfare effects and instead concentrate narrowly on whether vaccine sharing is good for the donating country rather than the recipient because self-interest is likely to be a strong motivation for embracing the cooperative policy. Roughly, is it in the self-interest of the US to share its scarce vaccine asset?

Figure 2 provides three examples, each connected to a different qualitative behavior of the system. In these simulations we focus on the US and on the COVAX AMC lower income countries (LIC) (for the list of countries see supplementary materials, section 3.3). We fix *R*_0_=2.5 and we keep the rest of the world with susceptibility equal to 35%, which means that these countries are above herd immunity levels and so they do not contribute significantly to the spread of the disease. We then consider an initial outbreak of 1000 new infections taking place in COVAX AMC LIC at time 0 and we follow the spread of the disease in the US under three scenarios. The baseline scenario considers the case of not having any extra doses of vaccine. The uncooperative scenario models the case in which 60 million extra doses are distributed in the US (we consider 2 doses of vaccine to be necessary for 95% efficacy). Finally, the cooperative scenario studies the case in which 60 million doses are given to COVAX AMC LIC (cooperative strategy). The first panel on the left considers the case in which both the US and COVAX AMC LIC countries have 35% of their population susceptible to COVID-19 before distributing the vaccine. As both countries have achieved herd immunity, we see how the impact on the US is negligible. The extra doses marginally reduce contagion, but to a degree that can be approximated to 0 (note the very small scale of the vertical axis). In the central panel of Figure 2 we see the case in which the US starts with a share of susceptible equal to 35%, while COVAX AMC LIC countries have 45% of their population at risk (below herd immunity). In this case, in COVAX AMC LIC countries the disease would spread to 5% of the population without extra doses of vaccine, and then begin to spread to the United States. When an outbreak takes place without additional vaccinations, the number of cases in the US peaks sharply, while it is suppressed by about 2/3rd when the additional vaccine doses are allocated domestically. However, allocating the additional doses cooperatively to the COVAX AMC LIC countries avoids this spillover all together by preventing the domestic outbreaks that would later spread to the US. In this case we therefore show that cooperation actions would allow for a reduction in the spread of the disease *in the US*. Finally, the panel on the right depicts a case in which both the US and the COVAX AMC LIC countries have relatively high rates of vulnerability and are thus below herd immunity. In this case, while allocating the doses to other countries delays the arrival of cases in the US, domestic allocation is more efficacious for protecting the domestic population because it allows the US to achieve herd immunity.

**Fig 2.**
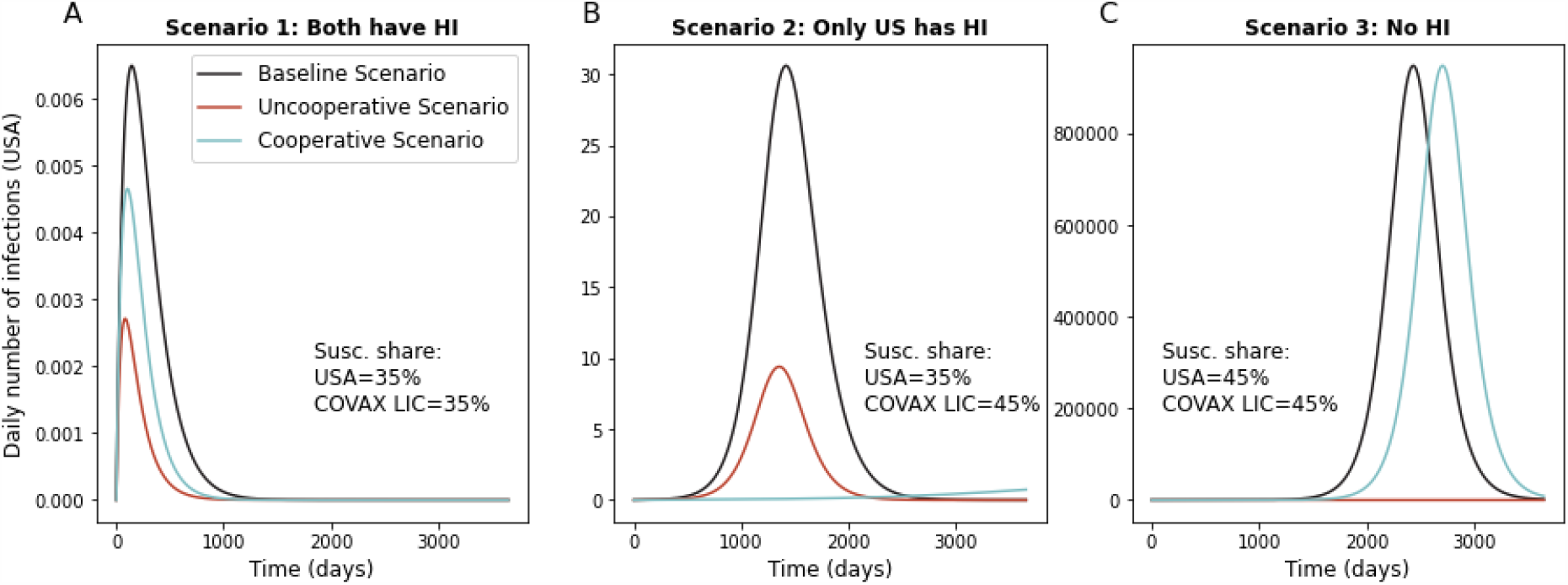
Daily number of infections in the US, comparison between Uncooperative and Cooperative scenarios. The figure compares between distributing 60mln extra doses in the USA (uncooperative scenario: red line) or in COVAX AMC LIC (cooperative scenario: light blue). In the baseline scenario (black line) the extra doses are not distributed. All panels consider *R*_0_ = 2.5 and the initial share of susceptible in the rest of the world equal to 35%. **(A)** The figure shows the daily number of infections in the US as estimated by the SIR model, when at time 0 the number of infected individuals equals 1000 in COVAX AMC LIC and is 0 everywhere else. The share of susceptible is 35% in both the US and in the COVAX AMC LIC. **(B)** As (A), but with initial shares of susceptible equal to 35% in the US and 45% in COVAX AMC LIC. **(C)** As (A), but with initial shares of susceptible equal to 45% in the US and 45% in COVAX AMC LIC.

Figure 3 extends the analysis done in Figure 2 to a wider set of combinations of susceptibility levels in the US and in the COVAX AMC LIC countries. The x-axis is the level of susceptibility in the US and the y-axis is the level of susceptibility in the COVAX AMC LIC countries. The plots display, for three different values of *R*_0_, level curves for the difference in cases between the two distribution strategies with 60 million vaccine doses: keeping them in the US or distributing them abroad. We show that there are a number of circumstances, those shown in blue in the plots, in which the US would be better off by using those doses abroad rather than internally when considering only the benchmark of domestic US cases. As Figure 3 shows, for a high enough share of susceptible in the US, the uncooperative strategy would guarantee a lower number of domestic infections. This is especially true in the dark shaded area, corresponding to the region where keeping the extra doses for the local population would allow the US to reach herd immunity. In blue, we highlight regions in which the cooperative strategy would lead to a smaller number of infections in the US. This is due to two factors. First, these are regions in which the US has already reached herd immunity so the marginal return to additional vaccinations is relatively low (see supplementary materials, section 3.1). Additionally, other countries are close enough to herd immunity that these additional doses allow them to achieve herd immunity and prevent sustained domestic outbreaks.

**Fig 3.**
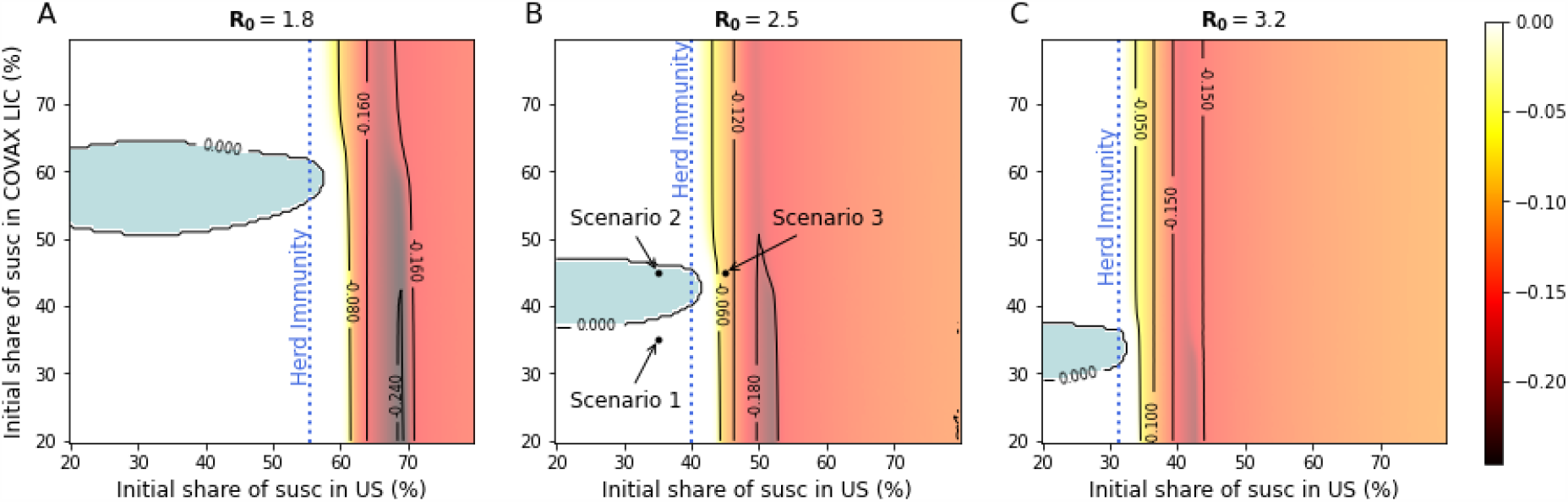
Share of individuals reached by the infection in the US, difference between uncooperative and cooperative scenarios. Difference between the share of infected in the US after 60 mln extra doses are distributed in the US (uncooperative scenario) and the share of infected in the US after the same number of doses is distributed in COVAX AMC LIC (cooperative scenario) for different values of *R*_0_ and susceptibility levels. Positive numbers (in blue) indicate a lower share of infected in the cooperative scenario. The shares of infected in the US are estimated using a SIR compartmental model and consider the whole evolution of contagions over the time span of 10 years. As initial condition, at time 0 we set the number of infected individuals equal to 1000 in COVAX AMC LIC and 0 everywhere else. (A) Difference in shares of infected, under the assumption that *R*_0_ = 1.8. (B) As (A), but assuming *R*_0_ = 2.5. (C) As (A), but assuming *R*_0_ = 3.2. Each point corresponds to a different combination of share of susceptible at time 0 in the USA and in the COVAX AMC LIC. In (B), Scenario 1 to 3 refer to panels (A) to (C) in Fig. 2.

## Conclusion

Overall, our results show that once the US reaches herd immunity level, it might be more beneficial for them to engage in a cooperative vaccination scenario rather than uncooperative scenario to prevent domestic infections. Our results build on the findings by Chinazzi et al.^15^ that show a cooperative strategy resulted in approximately twice the deaths averted globally but instead highlight that may be in a donor country’s self-interest to share vaccines once it achieves a certain vaccination threshold. Importantly, as vaccination becomes widespread in the US, such a distribution strategy will allow the US to keep its international borders open and avert economic losses inherent to the border closing.

Our analysis approaches the question of optimal vaccine distribution from a country’s self- interested perspective, something that has not been explored in prior work. The use of a simple but analytically solvable network SIR model allows us to make general predictions and compare many different scenarios. Model parameters are based on current understandings of SARS-CoV-2 and vaccine efficacy, which may change as more information becomes available. However, our model is robust to different specifications of these parameters. We also use travel data from the 2019 for network specifications. This does not account for reduced travel and travel restrictions in place during the pandemic. However, as our goal is to evaluate what is in a country’s self-interest, including economic interest, a pre-pandemic specification is more relevant for understanding how to maximize a country’s well-being.

## Data Availability

Data downloaded from International Civil Aviation Organization cannot be shared, but can be purchased from the provider.

https://doi.org/10.5281/zenodo.4432995

## Funding

PP and AM acknowledges support from the Italian Ministry of Education Progetti di Rilevante Interesse Nazionale (PRIN), respectively grant number 2017ELHNNJ and 20177BRJXS. AAM., EL, KW, SO acknowledge support from the Yale Institute for Global Health.

## Author contributions

TR and PP are joint primary authors. TR and PP designed the model. TR and MA collected data. TR PP, AM, MC analyzed the data. TR, PP, AM, MC, AAM, OS, EL, KW, GH, SB interpreted the results. TR, PP, AM, MC, AAM, GH wrote the manuscript. All authors edited the manuscript.

## Competing interests

None

## Ethics statement

No human subjects data was used or produced during this research.

## Data and code availability

All code will be made publicly available to download. Data on flight connections are restricted and can be downloaded from the provider’s website.

## Notes

### Competing Interest Statement

The authors have declared no competing interest.

### Author Declarations

No human subjects data was used or produced during this research.

