## Supplementary material for "National interest may require distributing COVID-19 vaccines to other countries": Methods, Supplementary Figures

†co-primary authors

**Methods**

Single country SIR model

We start from the standard SIR model (see *(16)* for a history of this model). The equations of the SIR model are

$$\frac{dS}{dt} = -\beta IS \quad (1)$$

$$\frac{dI}{dt} = \beta IS - \nu I \quad (2)$$

$$\frac{dR}{dt} = \nu I \quad (3)$$

where *S*, *I* and *R* are respectively the fraction of susceptible, infected and recovered people. $\beta$ and $\nu$ are respectively the infectiousness of and recovery rate from the disease. Note that $\beta$ are $\nu$ depend on policy choices that are, for instance, mask wearing, social distancing, quarantine measures, health care expenditures. Note also that, if there is a vaccination that guarantees temporary immunization, also the level $S_0$ of *S* that we start from is a policy choice: how many people to maintain vaccinated in steady state.

We call $R_0 = \beta/\nu$. The system, in a situation in which $I = 0$ and $R \geq 0$, is stable to perturbations if $S(t=0) < 1/R_0$, which is to say that $R > 1 - 1/R_0$. We assume $R_0 > 1$, to consider the interesting case where computations are not trivial, and we call $P_c = 1 - 1/R_0$ the threshold for *herd immunity*.

Starting form a stable situation, with $S = S_0$, we can compute the number of additional infection (leading eventually to recoveries) if there is an initial shock $I(t=0)$. Starting by dividing (2) by (1), We have:

$$\frac{dI}{dS} = -1 + \frac{1}{SR_0}$$

$$dI = \left(-1 + \frac{1}{SR_0}\right) dS \quad (4)$$

$$\int_0^\infty dI = \int_0^\infty \left(-1 + \frac{1}{SR_0}\right) dS$$

$$[I]_0^\infty = \left[-S + \frac{\log(S)}{R_0}\right]_0^\infty$$

$$I(t=0) = \phi(S(t=0)) - \phi(S_\infty)$$

where we have called $\phi(x) = x - \frac{\log(x)}{R_0}$.

So, how many additional people will get infected can be obtained (in general, non–analytically) from this $\phi(x)$ function.

The infection can spread to a limited part of the population (and not many) only if $\varphi$ is decreasing in $s_0$ (which is just the previous condition $S_0 < 1/R_0$, which means starting from a stable situations).

The multiplicative effect of a small shock, when starting from a stable steady, state can be computed as $1/\phi'$, which we define as $\delta = \frac{S(t=0)R_0}{1-S(t=0)R_0}$. This number is greater when we are close to the minimal herd immunity (at the limit $S(t = 0) \rightarrow 1/R_0$ it is infinite), and goes to 0 as $S(t = 0) \rightarrow 0$. This means that if the system is stable, but close to the minimal level of herd immunity, a very small shock can still propagate a lot. In this way, we relate the propagation of small shocks to the three policy variables that a country has: $\beta$, $v$ and $S(t = 0)$. $\delta$ is increasing in $\beta$ and $R_0$, and it is decreasing in $v$.

Note that $\delta$ can be obtained directly from (4), without taking the integrals. This is because we are just approximating linearly the effects of small shocks.

Network of countries

Consider a situation where there are $n$ connected countries. Each country $j$ has its own $R_{0,j} = \beta_j/\nu_j$ and its own $P_{c,j}$. All these variables are heterogeneous across countries because they depend on the medical and social policies that are chosen

Consider a network **G** where $g_{k,j}$ is the number of people moving from country $k$ visiting country $j$, divided by the population of country $j$. **G** is null in the diagonal.

In the network, equation (2) becomes:

$$\frac{dI_j}{dt} = \beta_j \left( I_j + \sum_k I_k g_{kj} \right) S_j - \nu_j I_j$$

In this case we can include the network effect in equation (4), obtaining:

$$dI_j = \left( -1 + \frac{1}{S_j R_{0,j}} - \sum_{k=1}^{n} g_{kj} \frac{I_k}{I_j} \right) dS_j \qquad (5)$$

Let us call **D** the diagonal matrix where the diagonal element $djj$ is $\frac{1}{S_j R_{0,j}}$. **I** is the identity matrix. Then, under the assumptions that for small initial shocks we have $\frac{I_k}{I_j} \simeq 1$, we can write equation (5) in vectorial form as

$$d\vec{I} = (-I + D - G)d\vec{S}$$

This can be inverted, to find with a linear approximation how many new infected (eventually recovered) will result from an initial shock *I(t=0)*. That is:[1]

[1] One way to interpret this expression is with a discrete time approach. The first–order effect of the contagion $d\vec{\imath}$ is $(\boldsymbol{D} - \boldsymbol{G})\boldsymbol{d\vec{\imath}}$: the diagonal element $d_{jj}$ accounts for recoveries, while $\boldsymbol{G}$ has all the network effects – we have a

$$d\vec{s} = (-\boldsymbol{I} + \boldsymbol{D} - \boldsymbol{G})^{-1} di$$

So, matrix $(-\mathbf{I} + \mathbf{D} - \mathbf{G})^{-1}$ tells us how the level of infection is from each country to any other country.

To have a clearer interpretation, let $\vec{m}$ be the inhabitants of each country and $Diag(\vec{m})$ be the diagonal matrix in which the diagonal consists of vector $\vec{m}$.

With this notation in mind, the matrix

$$Diag(\vec{m})(-\boldsymbol{I} + \boldsymbol{D} - \boldsymbol{G})^{-1} Diag(\vec{m})^{-1} \tag{6}$$

tells how many people in each column–country will be infected if one person is infected in row–country.

Measures of risk

We can start from the matrix expressed in (6) to identify some intuitive measures of risk for the countries. To begin, consider that if we set $d\vec{i}$ to a vector $\vec{1}$ of all 1's, we obtain a vector $\vec{c} = (-\boldsymbol{I} + \boldsymbol{D} - \boldsymbol{G})^{-1}\vec{1}$. A country $j$ that has $c_j$ relatively higher with respect to other countries, will be more at risk of propagation of the infection among its population.[2] So, this vector tell us which country is more susceptible to the propagation of the infection.

This approach allows us to study how a country can reduce its own $c_j$ playing at one side with the medical parameters $\beta_j$ (lockdown measures, policies on masks...), $\nu_j$ (better cures, isolation of infected people...) and $R(t=0)_{,j}$ (number of vaccinated people, once a vaccine is available) – all these factors affect matrix **D**; at the other side with **G** (possibly closing borders, or controlling people entering the country). Vector $\vec{c}$ includes all these aspects and takes into account the externalities between countries.

The problem with vector $\vec{c}$ is that it has not an immediate intuitive interpretation. Now, to express more intuitive measures, we need a vector $\vec{p}$ assigning the probability that a first positive infect appears in each country. We do this by obtaining two vectors.

First, we consider which country is more at risk.

$$Diag(\vec{m})(-\boldsymbol{I} + \boldsymbol{D} - \boldsymbol{G})^{-1} Diag(\vec{m})^{-1}\vec{p}$$

is a column vector that says how at risk each country is. Entry $i$ of this vector says, in expectation, how many people will become infected in country $i$ if a first person is infected in any of the countries.

---

minus sign because this is the decrease in susceptible people. But there is a second order effect in the second step, which is $(\boldsymbol{D} - \boldsymbol{G})^2 d\vec{\boldsymbol{i}}$; and then $(\boldsymbol{D} - \boldsymbol{G})^3 d\vec{\boldsymbol{i}}$, and so on...This amount exactly to

$$\sum_{t=1}^{\infty} (D - G)^t d\vec{i} = (-\boldsymbol{I} + \boldsymbol{D} - \boldsymbol{G})^{-1} d\vec{i}$$

This formula is equivalent to the formula for Bonacich centrality [17, 18].

[2] Note that, without network effects, this expression will give us $c_j = \delta_j = \frac{s_{0,j}\rho_j}{1 - s_{0,j}\rho_j}$, which was the result in isolation.

A simple candidate for $\vec{p}$ is $\vec{m}/(\vec{m}\cdot\vec{1})$, where $\vec{1}$ is a vector of all 1's and · is the vector multiplication. This assumes that the probability that an infection starts is a country is simply proportional to its population. Under this assumption, the previous vector is

$$Diag(\vec{p})(-\boldsymbol{I}+\boldsymbol{D}-\boldsymbol{G})^{-1} \tag{7}$$

We call this the **risk measure.**

Second, we consider which country is more dangerous.

$$\vec{1}'Diag(\vec{m})(-\boldsymbol{I}+\boldsymbol{D}-\boldsymbol{G})^{-1}Diag(\vec{m})^{-1} \tag{8}$$

is a row vector that says how dangerous each country is. Entry $i$ in this vector tells us how many infections will be caused by a person getting infected in country $i$. We call this the **danger measure**.[3] One could also change the formula in (8), so that the diagonal elements are removed from $(-\mathbf{I}+\mathbf{D}-\mathbf{G})^{-1}$: in this case we would have a **danger to others measure**, because we should not compute those that eventually become infected in the country where the disease originated.

These result are valid for small effects, that are obtained when all countries have herd immunity. To study the effects when a country is below herd immunity we must rely on simulations.

### Simulations

<u>The marginal effect of vaccinations</u>

We proceed by simulating the system described by equations (1), (2), and (3). The data on population were retrieved from The World Bank. We first simulate the model for the US, to study which fraction of the population would be eventually reached in case of an initial shock of 1000 people infected at time 0. We let the model run for 10 years and ignore the rest of the network. Figure S3 shows how the fraction of population eventually reached by the infection is a function of the initial share of susceptible. This analysis allows to study the reduction in the number of infections that can be obtained by reducing by 30 million the number of susceptible individuals in the country, this can be interpreted as the marginal effect of 60 million doses of vaccine. This number is function of the initial share of susceptible individuals. When the initial share of susceptible individuals is low, the reduction in the number of infections is close to 0, as the initial shock would propagate little across the population. The number grows and reaches a peak that corresponds to the herd immunity threshold. In this point the extra doses of vaccine would have the largest impact as they would allow the system to reach herd immunity. Figure S4 shows the same quantities for 2 million doses.

<u>Simulations with data on flights</u>

We proceed by simulating the system described by equations (1), (5), and (3) using data on flights for the year 2019. Data were obtained from ICAO and contain information on the number of passengers in international flights. In the network we therefore use countries as nodes. Links represent the average daily number of passengers going from one country to another. A graphical representation of this network is in Figure S1. Figure S2 shows the same network, where nodes sizes are proportional to the risk measure defined above.

[3] Note that, even if matrix $G$ is symmetric, risk measure from (7) and danger measure from (8) are different, because of the different population size of the countries.

We follow (*10, 19)* in setting $\gamma = 1/18$ that corresponds to an average duration of illness of 18 days. We simulate the model under various assumptions for the level of susceptible individuals in each country.[4] When considering countries belonging to COVAX AMC, we aggregate so that they appear as a single unit. To do that, we take the total population summing across all countries. For the network, we perform a similar operation, considering all flights that connect any of the COVAX AMC countries with any other country in the network.

<u>COVAX AMC countries</u>

COVAX AMC Lower Income Countries: Afghanistan, Benin, Burkina Faso, Burundi, Central African Republic, Chad, Dem. Rep. of the Congo, Eritrea, Ethiopia, Gambia, Guinea, Guinea-Bissau, Haiti, Liberia, Madagascar, Malawi, Mali, Mozambique, Nepal, Niger, Rwanda, Sierra Leone, Somalia, South Sudan, Syrian Arab Republic, Tajikistan, Togo, Uganda, United Republic of Tanzania, Yemen Rep.
COVAX AMC Lower and Middle Income Countries: Afghanistan, Algeria, Angola, Bangladesh, Benin, Bhutan, Bolivia, Burkina Faso, Burundi, Cabo Verde, Cambodia, Cameroon, Central African Republic, Chad, Comoros, Congo, Cote d'Ivoire, Dem. Rep. of the Congo, Djibouti, Egypt, El Salvador, Eritrea, Eswatini, Ethiopia, Gambia, Ghana, Guinea, Guinea-Bissau, Haiti, Honduras, India, Indonesia, Kenya, Kiribati, Kyrgyzstan, Lao PDR, Lesotho, Liberia, Madagascar, Malawi, Mali, Mauritania, Micronesia Fed. Sts., Mongolia, Morocco, Mozambique, Myanmar, Nepal, Nicaragua, Niger, Nigeria, Pakistan, Papua New Guinea, Philippines, Rwanda, Sao Tome and Principe, Senegal, Sierra Leone, Solomon Islands, Somalia, South Sudan, Sri Lanka, Sudan, Syrian Arab Republic, Tajikistan, Timor-Leste, Togo, Tunisia, Uganda, Ukraine, United Republic of Tanzania, Uzbekistan, Vanuatu, Viet nam, West Bank and Gaza, Yemen, Rep., Zambia, Zimbabwe.

### Methods References


1. J. A. P. Heesterbeek. A brief history of $R_0$ and a recipe for its calculation. *Acta biotheoretica*. **50,** 189–204 (2002).
2. P. Bonacich. Power and centrality: A family of measures. *American journal of sociology*. **92** 1170–1182 (1987).
3. V. Dequiedt and Y. Zenou. Local and consistent centrality measures in parameterized networks. *Mathematical Social Sciences*. **88**, 28–36 (2017).
4. H. Wang, Z. Wang, Y. Dong, R. Chang,C. Xu, Y. Xu, X. Yu, S. Zhang, L.Tsamlag, M. Shang, J. Huang, Y. Wang, G. Xu, T. Shen, X. Zhang, Y. Cai Phase-adjusted estimation of the number of Coronavirus Disease 2019 cases in Wuhan, China. *Cell Discov* **6,** 10 (2020).


4 To solve the system of differential equations we use SciPy integrate.solve ivp solver, using Adams/BDF method.

**Extended data figure**

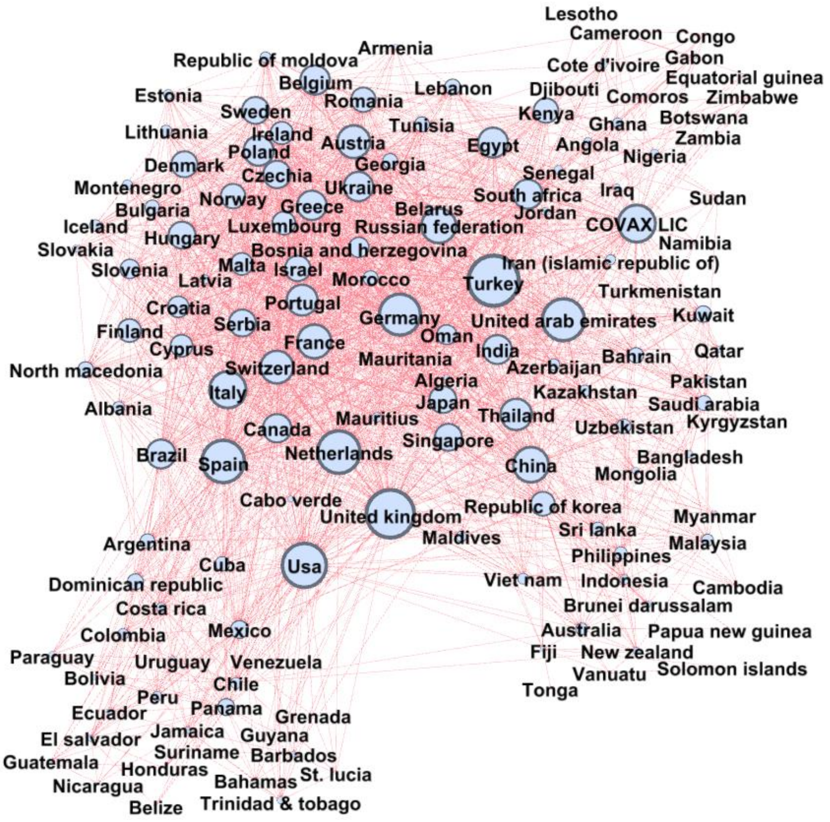


**Fig S1. Network of Flights**. Nodes are countries and the size depends on the total inflow of passengers reaching each country. A direct link denotes the existence of a direct commercial flight between the two countries.

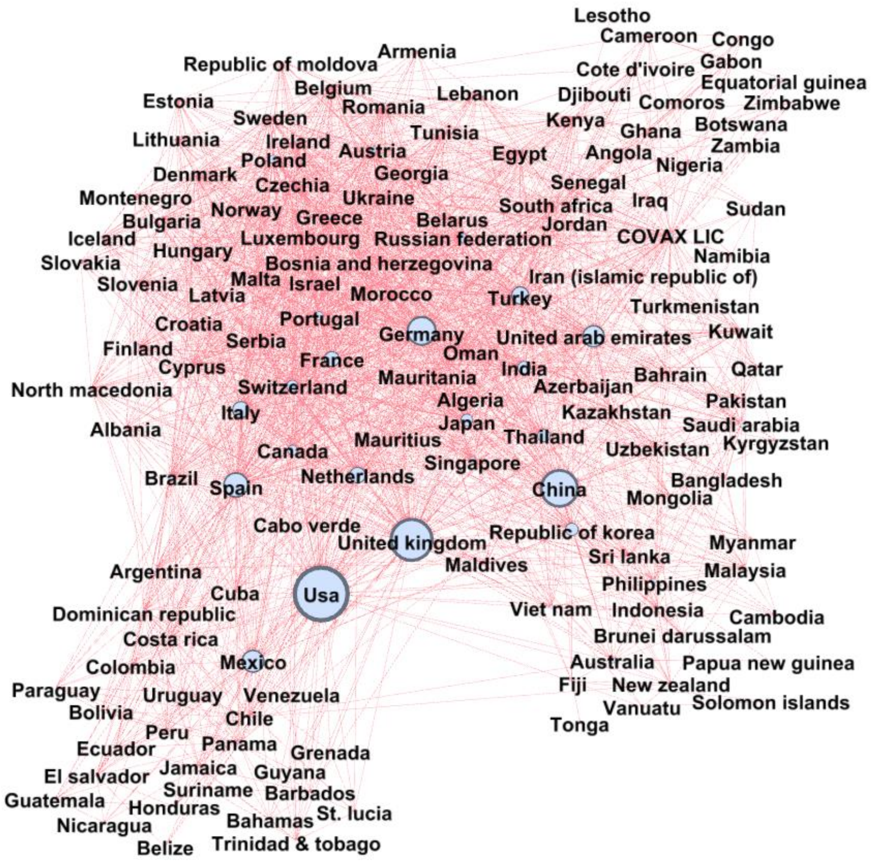


**Fig S2. Network of Flights**. Nodes are countries and the size are proportional to the risk measure, calculated assigning a share of susceptible equal to 35% to every country. A direct link denotes the existence of a direct commercial flight between the two countries.

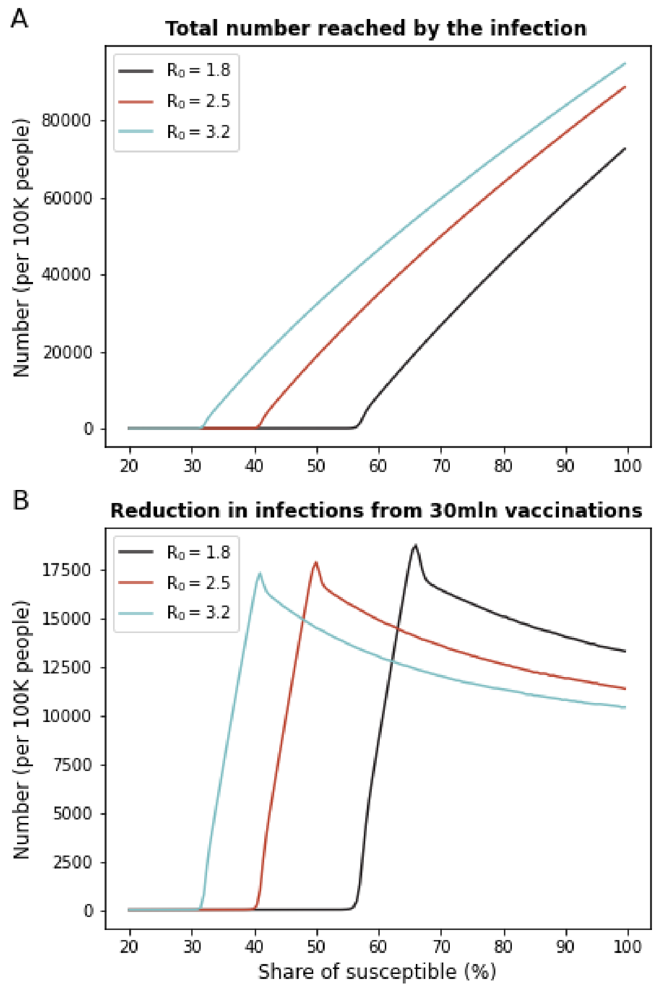


**Fig S3. Marginal effects of additional vaccinations. (A)** The figure shows the number of people eventually reached by the infection after 1000 individuals get infected at time 0. The number is expressed as a function of the initial share of susceptible individuals. **(B)** The figure shows the reduction in the number of infections that would be determined by 60 million additional doses (30 million immune individuals) done before the start of the contagion at time 0. This number varies as it depends on the spread of the disease determined by the share of susceptible individuals.

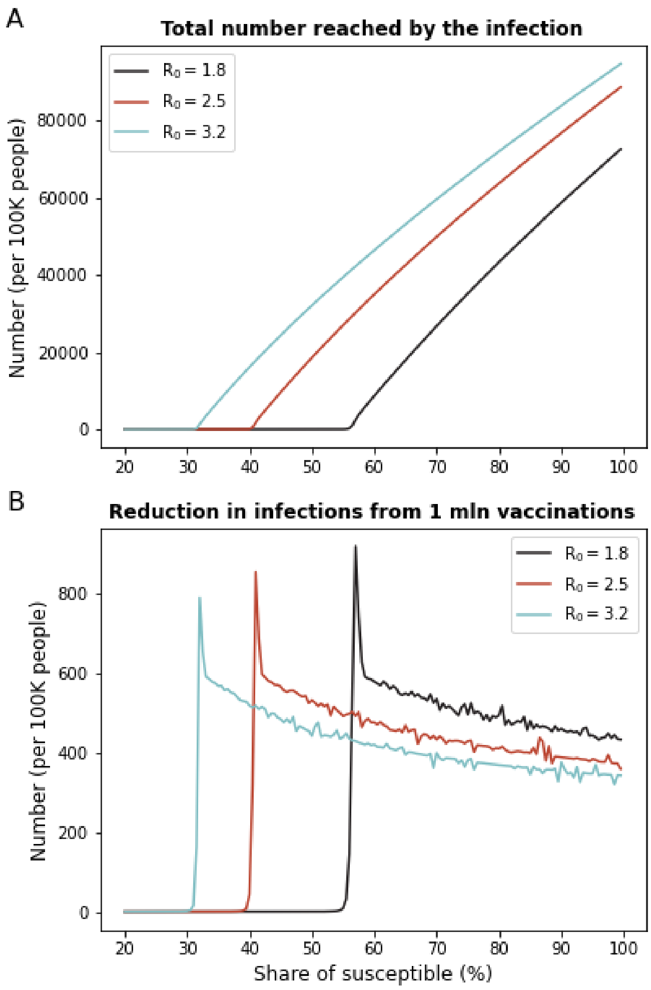


**Fig S4. Marginal effects of additional vaccinations. (A)** The figure shows the number of people eventually reached by the infection after 1000 individuals get infected at time 0. The number is expressed as a function of the initial share of susceptible individuals. **(B)** The figure shows the reduction in the number of infections that would be determined by 2 million additional doses (1 million immune individuals) done before the start of the contagion at time 0. This number varies as it depends on the spread of the disease determined by the share of susceptible individuals.

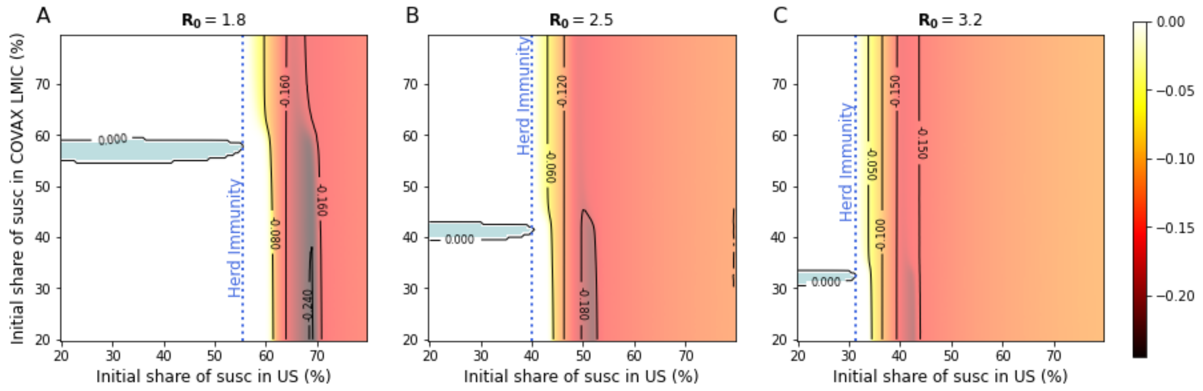


**Fig S5.** Share of individuals reached by the infection in the US, difference between uncooperative and cooperative scenarios. Difference between the share of infected in the US after 60mln extra doses are distributed in the US (uncooperative scenario) and the share of infected in the US after the same number of doses is distributed in COVAX AMC lower and middle income countries (cooperative scenario) for different values of $R_0$ and susceptibility levels. Positive numbers (in blue) indicate a lower share of infected in the cooperative scenario. The shares of infected in the US are estimated using a SIR compartmental model and consider the whole evolution of contagions over the time span of 10 years. As initial condition, at time 0 we set the number of infected individuals equal to 1000 in COVAX AMC lower and middle income countries and 0 everywhere else. (A) Difference in shares of infected, under the assumption that $R_0$ = 1.8. (B) As (A), but assuming $R_0$ = 2.5. Each point corresponds to a different combination of share of susceptible at time 0 in the US and in the COVAX AMC lower and middle income countries. (C) As (A), but assuming $R_0$ = 3.2.
